# Procedural Volume and Outcomes in Lower Extremity Peripheral Vascular Interventions: Insights from the National Cardiovascular Data Registry Peripheral Vascular Intervention Registry

**DOI:** 10.1101/2024.05.30.24308245

**Authors:** Zainali Chunawala, Arman Qamar, Kevin Kennedy, Neil Keshwani, Ambarish Pandey, Dharam J. Kumbhani, Cheong Jun Lee, Gregory Mishkel, Eric Secemsky, Deepak L. Bhatt

## Abstract

**Background:** Lower extremity peripheral vascular interventions (PVI) are increasingly utilized for treatment of peripheral artery disease (PAD). However, the relationship between hospital- and operator-volume of PVI with in-hospital major adverse limb events (MALE) and major adverse cardiovascular events (MACE) is not well established.

**Methods:** We utilized the National Cardiovascular Data Registry PVI registry that included procedural data from 4/1/2014 – 9/3/2020 and assessed in-hospital MALE and MACE during hospitalization for PVI. Generalized linear mixed models were used to assess the volume-outcome relationships after adjustment of relevant covariates.

**Results:** Between 2014 to 2020, a total of 60,834 PVI procedures were performed at 97 hospitals by 555 operators. In the adjusted analysis, there was no significant association between tertile of hospital volume and MALE (highest vs lowest-volume: OR 0.89, 95% CI 0.73–1.08, p=0.24) or tertile of hospital-volume and MACE (highest vs lowest-volume: OR 1.26, 95% CI 0.98 – 1.62, p=0.066). Notably, undergoing PVI with high volume operators was associated with lower odds of in-hospital MALE (highest vs lowest-volume: OR 0.72, 95% CI 0.54–0.94, p=0.017). However, no association was observed between operator-volume tertiles and MACE (highest vs lowest-volume: OR 0.71, 95% CI 0.48–1.04, p=0.082).

**Conclusions:** In contemporary practice, a strong relationship was observed between operator volume and adjusted PVI outcomes. Highest volume operators had lower rates of in-hospital MALE compared with lowest volume operators. However, there was no observed relationship between tertiles of hospital volume of PVI and associated outcomes.

**Clinical Perspectives:** *What is New?:*

- Procedural volume has been defined as a surrogate to define quality of care in many cardiovascular settings.
- In patients undergoing peripheral vascular intervention, highest volume operators were associated with lower incidence of in-hospital major adverse limb events.
- However, there was no observed association between hospital volume of peripheral vascular interventions and outcomes.

*Clinical Implications:*

- Operator-volume may be a valuable surrogate to assess peripheral vascular interventions quality across different centers and could potentially outweigh the significance of institutional volume in the context of the procedural safety and outcomes.

## Introduction

Lower extremity peripheral artery disease (PAD) has emerged as a critical health concern, causing substantial morbidity and mortality, with an estimated 12.5 million Americans affected.^1–3^ The cost associated with hospitalization and revascularization procedures for patients with PAD continues to rise, intensifying the economic burden.^4–6^ Lower extremity peripheral vascular interventions (PVI) have expanded significantly and gained prominence as one of the primary approaches to revascularization in patients with PAD.^7–9^ However, given the increasing prevalence of PAD, the expanding utilization of PVI as a treatment modality, and its significant cost burden, ensuring optimal quality of care of PAD patients undergoing PVI is of principle importance.

Previously, the role of procedural or surgical volume, both at the individual operator and site levels, has been investigated as a surrogate to define quality of care in many cardiovascular settings.^10–14^ Despite the apparent intuitiveness and ease of collecting volume as a metric, there exist a significant degree of heterogeneity in PVI procedural utilization nationwide arising from differences in operator specialty, training requirements and procedural location. Furthermore, the relationship between procedural volume and patient outcomes in the context of PVI remains largely unknown. Accordingly, the objective of our study was to investigate the association between the volume of PVI procedures conducted by hospitals and individual operators and the occurrence of in-hospital major adverse limb events (MALE) and major adverse cardiovascular events (MACE).

## Methods

### Study Population & Cohort Derivation

The National Cardiovascular Data Registry Peripheral Vascular Intervention (NCDR PVI) Registry is a large, nationwide, voluntary, continuous quality improvement registry of all patients undergoing percutaneous vascular interventions. The NCDR PVI Registry contains detailed information on patient demographics, clinical characteristics and comorbidities, procedural information, and outcomes. Additionally, the registry captures procedure operator and facility characteristics. Details regarding data collection and standardization and registry structure and governance have been published previously.^15^ Data managers at each center supervise data collection, and data are submitted securely through a web-based data collection tool. Waiver for informed consent was granted by the Advarra Institutional Review Board. All PVI procedures (N = 60,834) from April 1, 2014 to September 30, 2020 were included in the analysis at 97 sites by 555 operators.

### Outcomes

The primary outcome were in-hospital major adverse limb events (MALE) and major adverse cardiovascular events (MACE) during the hospitalization. MALE was defined as a loss of patency of the revascularization, repeat intervention, or a major amputation of the revascularized limb. MACE was defined as a cerebrovascular event, myocardial infarction (MI), or death. Other key secondary outcomes included individual components of the composite endpoints, length of stay, unexpected intubation, bleeding event within 72 hours, or blood transfusion.

### Clinical Characteristics

Baseline patient characteristics included age at the time of PVI, sex, race, and other sociodemographic factors, clinical comorbidities and characteristics including hypertension, dyslipidemia, diabetes mellitus, end-stage renal disease (ESRD) on dialysis, coronary artery disease (CAD), severe or very severe lung disease, prior myocardial infarction (MI), cardiomyopathy or left ventricular systolic dysfunction, prior heart failure (HF), prior percutaneous coronary intervention (PCI), prior coronary artery bypass grafting (CABG), prior valve surgery, tobacco use, and family history of premature CAD. Clinical information on stage of PAD at presentation and maximum claudication distance were included. Procedural status (elective, urgent, or emergent), procedure indication, procedure location, and procedural information such as procedural vital signs, sedation, medication administration, contrast volume, and fluoroscopy time were included.

The annualized counts of hospital and operator procedure volumes were determined by dividing the cumulative number of PVI procedures performed at the hospital or by individual operator throughout the study duration by the number of months between the first and last procedure, then multiplying by 12 months. In instances where multiple operators participated in a procedure, each operator received credit for their contribution to the procedure.

### Statistical Analysis

Annualized hospital volume of PVI procedures was analyzed both as a categorical variables sorted into tertiles of either operator or site volume. Baseline and clinical characteristics were presented as frequencies (percentages) and continuous variables presented as mean (standard deviation) or median (interquartile range). Procedural characteristics and procedural outcomes were described across these tertiles. Categorical variables were compared using Mantel-Haenszel trend test and categorical variables compared using the linear trend test.

The primary analysis examined the association between tertile of hospital procedural volume and post-procedural MALE and MACE. The secondary analysis examined the association between tertile of operator procedural volume and post-procedural MALE and MACE. Generalized linear mixed models were developed to assess the volume-outcome relationships, with marginal estimates reported. Relationships were plotted as curves for annualized hospital procedural volume and outcomes and for annualized operator procedural volume and outcomes. Analyses were repeated following adjustment of relevant covariates including age, gender, race, history of hypertension, dyslipidemia, diabetes mellitus, peripheral artery disease, coronary artery disease, heart failure, lung disease, currently dialysis, tobacco use and family history of CAD, CLI and ALI. Further adjustment for operator volume was included in the assessment of hospital procedural volume and clinical outcome relationship. All analysis were performed with SAS version 9.4 (SAS institute, Cary, North Carolina).

## Results

### Hospital and Operator Procedural Volumes

Between Quarter 2, 2014 and Quarter 3, 2020 a total of 60,834 PVI procedures were performed at 97 hospitals by 555 operators. Annualized PVI hospital procedural volume ranged from 4 to 907 (median 134) procedures, while annualized operator procedural volume ranged from 4 to 256 (median 19.3) procedures. The spectrum of annualized hospital and procedural volume is shown in **Figure 1**. The overall procedural volume increased steadily until late 2019, which was followed by procedural volume decline through 2020. Trends in number of PVI procedures over the study period are shown in **Figure 2**.

**Figure 1.**
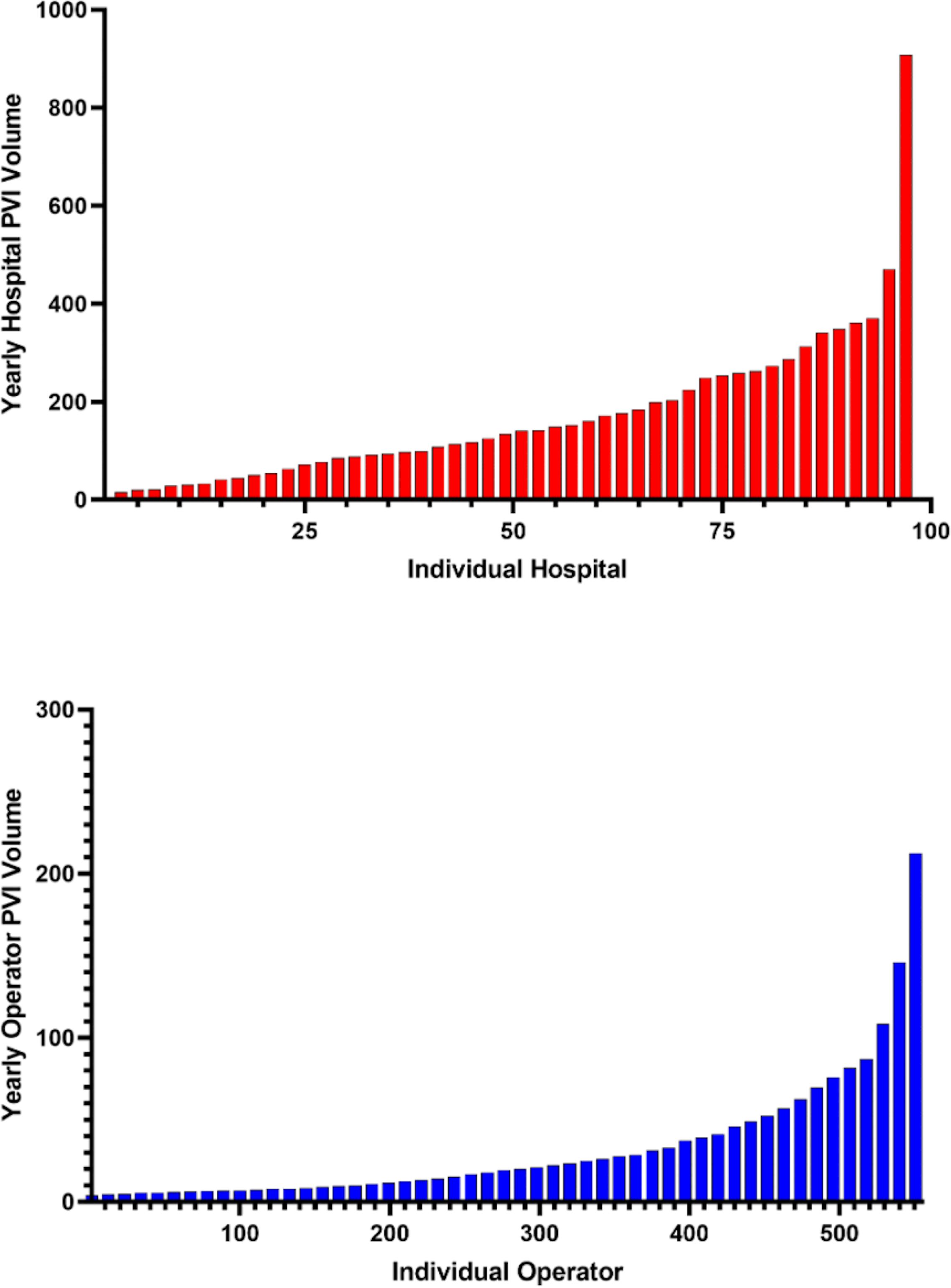
Annualized hospital and operator volume of peripheral vascular intervention procedures. Legend: A. Annualized PVI hospital procedural volume IQR: 4 to 907 (median = 134) B. Annualized PVI operator procedural volume IQR: 4 to 256 (median = 19.3)

**Figure 2.**
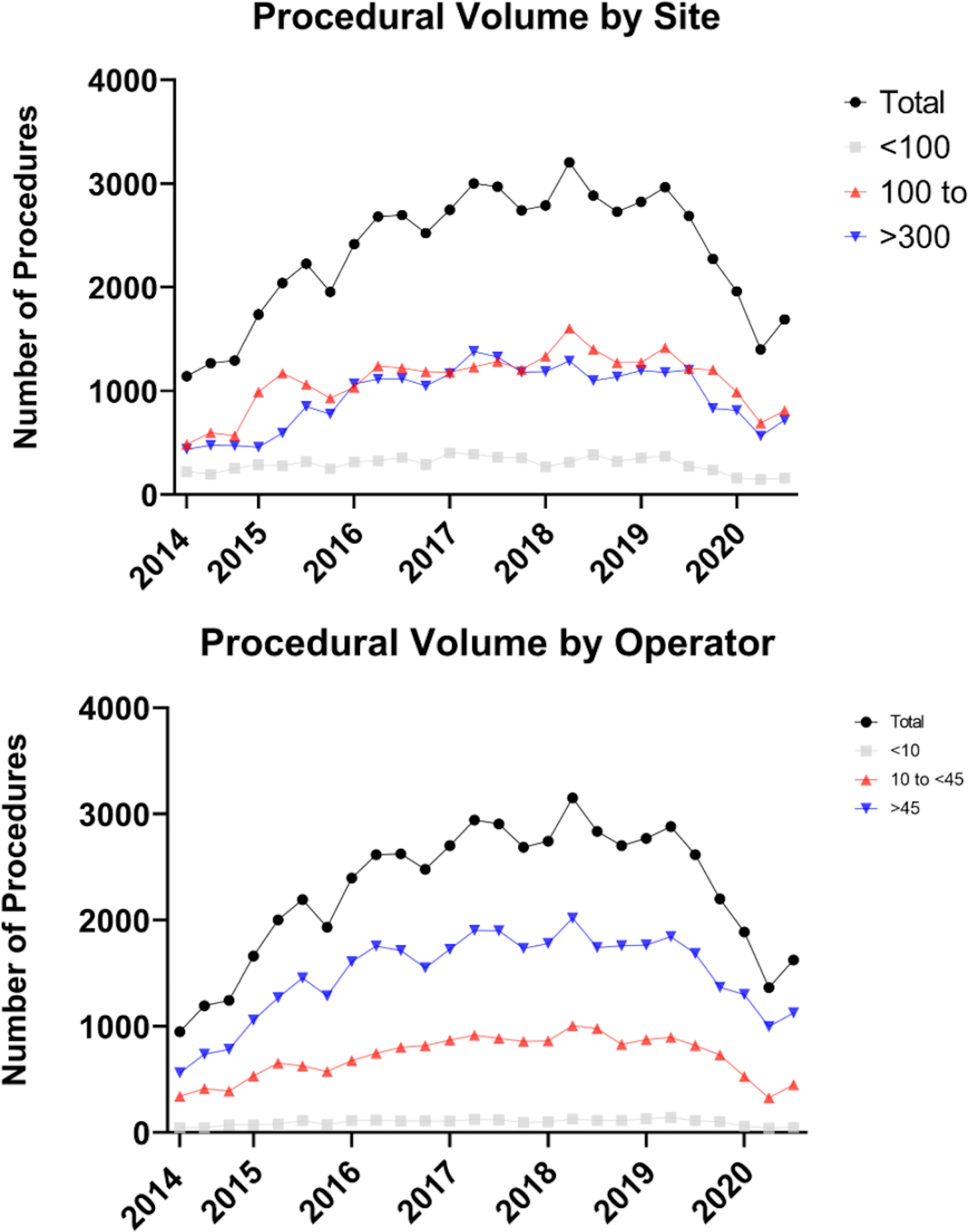
Trends in peripheral vascular interventions over the study period.

### Patient and Procedural Characteristics

Mean age of all patients who underwent PVI was 68.9 +/- 11.2 (80.1% White, 40.5% female). There was an overall high prevalence of comorbidities such as hypertension (90.1%), coronary artery disease (50.3%), and current or former tobacco use (76.8%). Of total procedures, 75.4% were classified as elective, 22.5% classified as urgent, and 2.1% were emergent. Most procedures were for typical claudication (42.7%) and critical limb ischemia (45.9%), with less frequent treatment of acute limb ischemia (5.9%). Overall, 81.6% of procedures took place in a catheterization laboratory, 6.4% in an operating room, and 12.0% in an interventional radiology suite.

Across tertiles of hospital-volume of PVI, there was no significant difference in age. (**Table 1**) Highest-volume centers cared for greater percentage of Black (23.1% vs 12.3%, p<0.001) patients and lower percentage of White patients (74.2% vs 85.1%, p<0.001). There was a significantly greater prevalence of comorbidities such as hypertension, dyslipidemia, prior MI, and prior HF among highest-volume centers. Lastly, greater percentage of procedures took place in an operating room (8.3% vs 3.2%, p<0.001) and fewer procedures in the catheterization laboratory (82.9% vs 93.8%, p<0.001) among the highest volume centers compared with lowest volume centers.

**Table 1.**
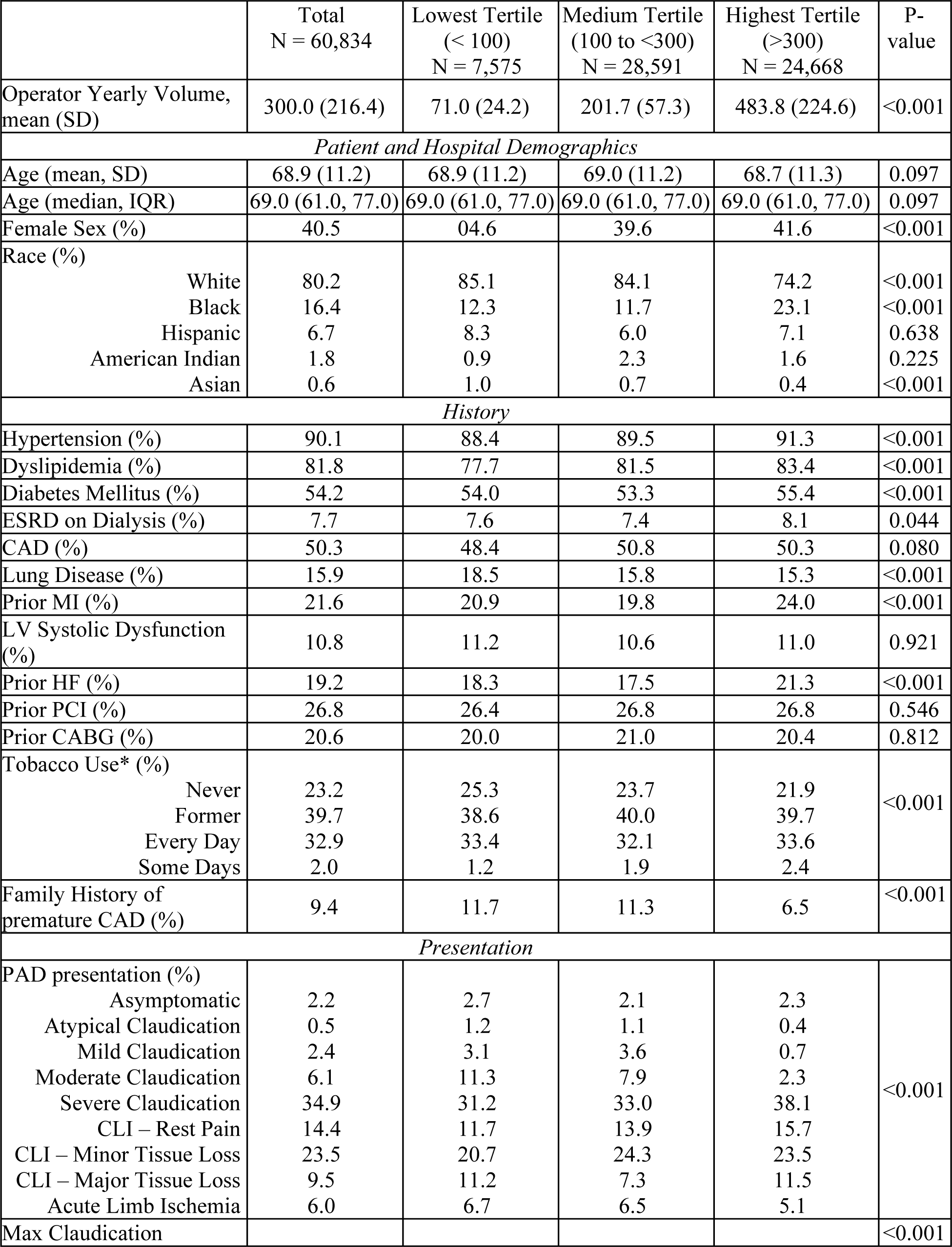
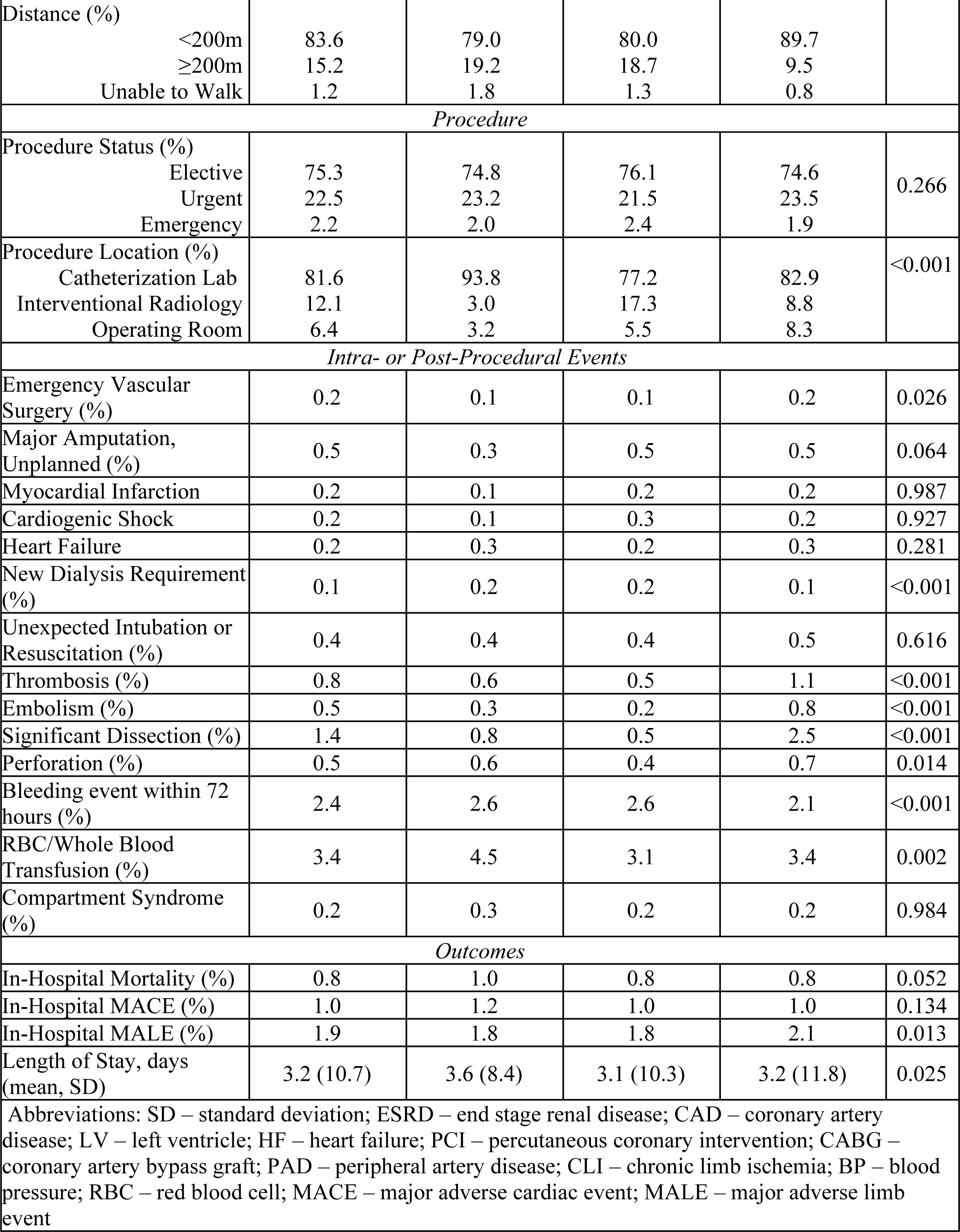
Baseline characteristics and clinical outcomes across tertiles of hospital volume of PVI.

Across tertiles of operator-volume of PVI, highest-volume operators cared for lower percentage of Black (16.9% vs 17.4%, p = 0.006) and Hispanic patients (6.4% vs 9.1%, P < 0.001) without significant differences in patient comorbidities or procedure location (**Table 2**). Operators with the highest volume cared for greater percentage of patients with critical limb ischemia (50.5% vs 34.6%, p<0.001) and fewer percentage of patients with typical claudication (41.3% vs 54.4%, p<0.001) as compared with lowest-volume operators.

**Table 2.** Baseline characteristics and clinical outcomes across tertiles of operator volume of PVI.

|  | Total<br>N = 59,294 | Lowest Tertile<br>(<br>< 10)<br>N = 2,472 | Medium Tertile<br>(10 to <45)<br>N = 18,409 | Highest Tertile<br>(>45)<br>N = 38,413 | P-value |
| --- | --- | --- | --- | --- | --- |
| Operator Yearly Volume,<br>mean (SD) | 83.3 (66.0) | 3.1 (5.8) | 3.5 (8.9) | 3.1 (11.8) | 0.862 |
| Patient and Hospital Demographics |  |  |  |  |  |
| Age (mean, SD) | 68.9 (11.2) | 69.4 (10.6) | 68.6 (11.2) | 69.0 (11.2) | 0.073 |
| Age (median, IQR) | 69.0 (61.0, 77.0) | 70.0 (62.0, 77.0) | 69.0 (61.0, 77.0) | 69.0 (61.0, 77.0) | 0.073 |
| Female Sex (%) | 40.5 | 37.8 | 40.3 | 40.7 | 0.016 |
| Race (%) |  |  |  |  | <0.001 |
| White | 80.1 | 78.9 | 81.9 | 79.3 |  |
| Black | 16.5 | 17.4 | 15.4 | 16.9 |  |
| Hispanic | 6.6 | 9.1 | 6.8 | 6.4 |  |
| American Indian | 1.9 | 1.3 | 0.9 | 2.4 |  |
| Asian | 0.6 | 1.0 | 1.0 | 0.4 |  |
| History |  |  |  |  |  |
| Hypertension (%) | 90.1 | 90.2 | 89.3 | 90.5 | <0.001 |
| Dyslipidemia (%) | 81.9 | 81.2 | 80.5 | 82.6 | <0.001 |
| Diabetes Mellitus (%) | 54.2 | 53.7 | 53.3 | 54.7 | 0.003 |
| ESRD on Dialysis (%) | 7.7 | 6.4 | 7.4 | 8.0 | <0.001 |
| CAD (%) | 50.3 | 55.3 | 52.5 | 49.0 | <0.001 |
| Lung Disease (%) | 15.9 | 16.1 | 15.6 | 16.0 | 0.444 |
| Prior MI (%) | 21.6 | 21.9 | 22.9 | 21.0 | <0.001 |
| LV Systolic Dysfunction (%) | 10.8 | 11.0 | 11.8 | 10.3 | <0.001 |
| Prior HF (%) | 19.2 | 19.2 | 20.3 | 18.6 | <0.001 |
| Prior PCI (%) | 26.8 | 30.3 | 28.5 | 25.8 | <0.001 |
| Prior CABG (%) | 20.6 | 23.0 | 22.0 | 19.8 | <0.001 |
| Tobacco Use* (%) |  |  |  |  | 0.002 |
| Never | 23.1 | 26.2 | 22.2 | 23.4 |  |
| Former | 39.8 | 40.3 | 41.5 | 38.9 |  |
| Every Day | 32.9 | 28.7 | 32.6 | 33.3 |  |
| Some Days | 2.0 | 2.2 | 1.9 | 2.1 |  |
| Family History of premature CAD (%) | 9.4 | 9.6 | 8.7 | 9.7 | 0.004 |
| Presentation |  |  |  |  |  |
| PAD presentation (%) |  |  |  |  | <0.001 |
| Asymptomatic | 2.1 | 3.0 | 2.6 | 1.9 |  |
| Atypical Claudication | 0.8 | 1.3 | 1.0 | 0.7 |  |
| Mild Claudication | 2.3 | 3.8 | 2.7 | 2.1 |  |
| Moderate Claudication | 6.0 | 12.2 | 7.1 | 5.1 |  |
| Severe Claudication | 35.0 | 38.4 | 36.4 | 34.1 |  |
| CLI – Rest Pain | 14.4 | 10.1 | 13.7 | 14.9 |  |
| CLI – Minor Tissue Loss | 23.6 | 16.7 | 20.2 | 25.7 |  |
| CLI – Major Tissue Loss | 9.6 | 7.8 | 9.2 | 9.9 |  |
| Acute Limb Ischemia | 5.9 | 5.7 | 6.8 | 5.5 |  |
| Max Claudication Distance |  |  |  |  | <0.001 |
| (%) |  |  |  |  |  |
| <200m | 83.8 | 80.3 | 83.0 | 84.6 |  |
| ≥200m | 15.0 | 18.3 | 15.37 | 14.3 |  |
| Unable to Walk | 1.2 | 1.4 | 1.3 | 1.1 |  |
| Procedure |  |  |  |  |  |
| Procedure Status (%) |  |  |  |  |  |
| Elective | 75.4 | 75.6 | 75.4 | 75.4 | 0.149 |
| Urgent | 22.5 | 22.0 | 22.0 | 22.7 |  |
| Emergency | 2.1 | 2.4 | 2.6 | 1.9 |  |
| Procedure Location (%) |  |  |  |  |  |
| Catheterization Laboratory | 81.6 | 83.6 | 80.3 | 82.0 | <0.001 |
| Interventional Radiology | 12.0 | 13.7 | 11.3 | 12.2 |  |
| Operating Room | 6.4 | 2.7 | 8.4 | 5.7 |  |
| Intra- or Post-Procedural Events |  |  |  |  |  |
| Emergency Vascular Surgery (%) | 0.2 | 0.1 | 0.2 | 0.2 | 0.994 |
| Major Amputation, unplanned (%) | 0.5 | 0.6 | 0.4 | 0.5 | 0.515 |
| Myocardial Infarction | 0.2 | 0.1 | 0.2 | 0.2 | 0.465 |
| Cardiogenic Shock | 0.2 | 0.2 | 0.2 | 0.3 | 0.159 |
| Heart Failure | 0.2 | 0.3 | 0.3 | 0.2 | 0.364 |
| New Dialysis Requirement (%) | 0.1 | 0.1 | 0.1 | 0.2 | 0.390 |
| Unexpected Intubation or Resuscitation (%) | 0.4 | 0.5 | 0.4 | 0.5 | 0.756 |
| Thrombosis (%) | 0.8 | 0.4 | 0.6 | 0.8 | 0.003 |
| Embolism (%) | 0.5 | 0.3 | 0.3 | 0.6 | <0.001 |
| Significant Dissection (%) | 1.4 | 0.5 | 0.9 | 1.7 | <0.001 |
| Perforation (%) | 0.5 | 0.6 | 0.4 | 0.6 | 0.017 |
| Bleeding event within 72 hours (%) | 2.4 | 3.1 | 2.4 | 2.3 | 0.029 |
| RBC/Whole Blood Transfusion (%) | 3.3 | 3.4 | 3.5 | 3.2 | 0.051 |
| Compartment Syndrome (%) | 0.2 | 0.1 | 0.3 | 0.2 | 0.173 |
| Outcomes |  |  |  |  |  |
| In-Hospital Mortality (%) | 0.8 | 0.9 | 0.9 | 0.7 | 0.017 |
| In-Hospital MACE (%) | 1.0 | 1.2 | 1.2 | 1.0 | 0.014 |
| In-Hospital MALE (%) | 1.9 | 2.3 | 1.8 | 1.9 | 0.842 |
| Length of Stay, days (mean, SD) | 3.2 (10.8) | 3.1 (5.8) | 3.5 (8.9) | 3.1 (11.8) | 0.862 |
| Abbreviations: SD – standard deviation; ESRD – end stage renal disease; CAD – coronary artery disease; LV – left ventricle; HF – heart failure; PCI – percutaneous coronary intervention; CABG – coronary artery bypass graft; PAD – peripheral artery disease; CLI – chronic limb ischemia; BP – blood pressure; RBC – red blood cell; MACE – major adverse cardiac event; MALE – major adverse limb event |  |  |  |  |  |

### Procedural Outcomes and Association with Volume

Highest volume centers had higher rates of arterial thrombosis (1.1% vs 0.6%, p<0.001), embolism (0.8% vs 0.3%, p<0.001), major dissection (2.5% vs 0.8%, p<0.001) and lower rates of bleeding events (2.1% vs 2.6%, p<0.001) or blood transfusion (3.4% vs 4.5%, p<0.001) as compared with lowest-volume centers. Overall, the rates of in-hospital outcomes were 1.9% for MALE, 1.0% for MACE and 0.8% for all-cause mortality. The incidence of MALE was greater in highest-volume centers compared with lowest volume centers (2.1% vs 1.8%, p = 0.013).

However, there were no significant difference observed in in-hospital MACE or in-hospital mortality across tertiles of hospital-volume. In the adjusted analysis, there was no association observed between tertile of hospital volume and MALE (highest vs lowest-volume: OR 0.89, 95% CI 0.73 – 1.08, p = 0.239) and tertile of hospital-volume and MACE (highest-vs lowest-volume: OR 1.26, 95% CI 0.98 – 1.62, p = 0.066) (**Figure 3**).

**Figure 3.**
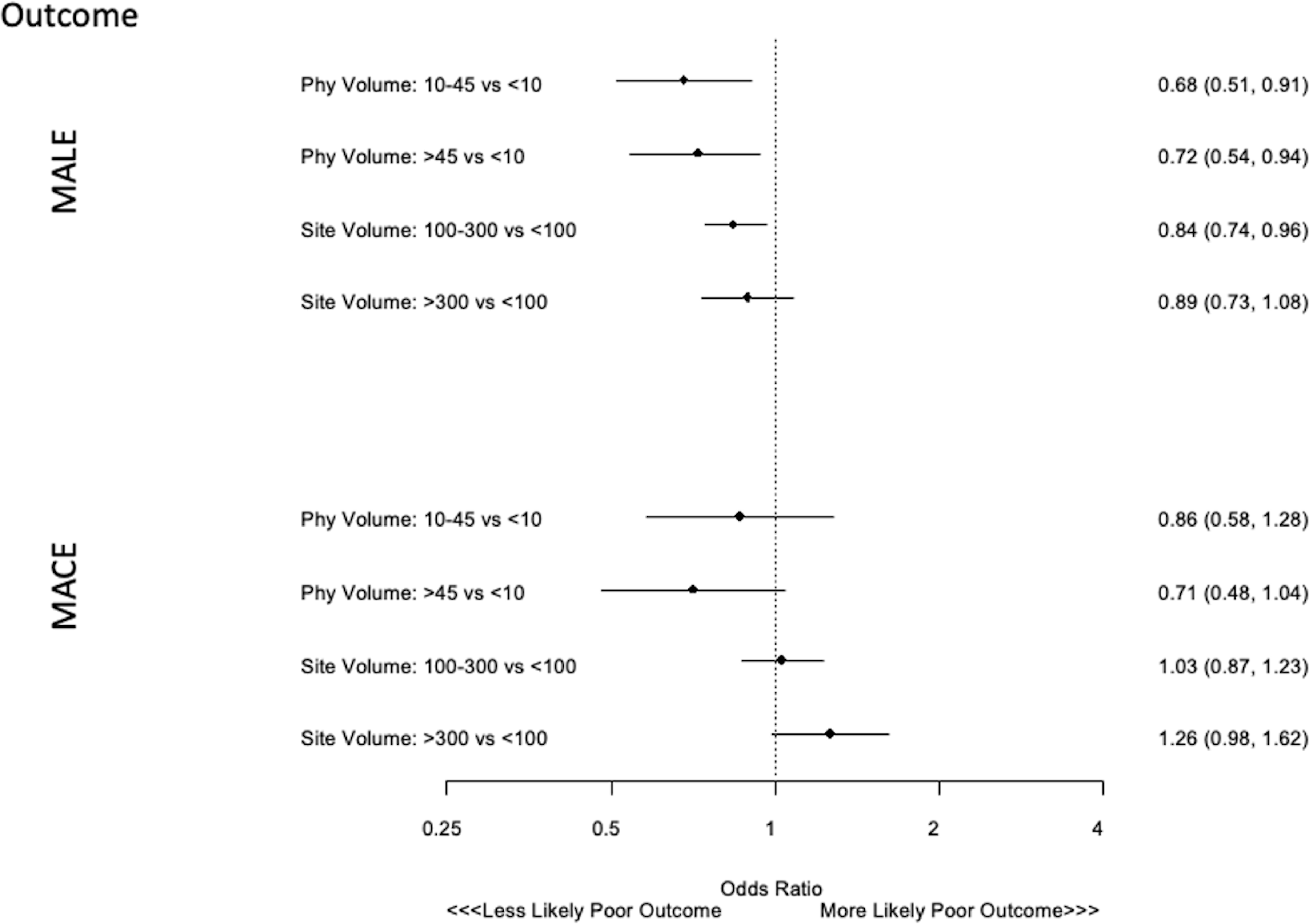
Association of tertile of operator and hospital volume with MALE and MACE. Legend: Model adjusted for covariates including age, gender, race, history of hypertension, dyslipidemia, diabetes mellitus, peripheral artery disease, coronary artery disease, heart failure, lung disease, currently dialysis, tobacco use and family history of CAD, CLI and ALI. MALE indicates major adverse limb events; MACE, major adverse cardiovascular events.

Highest volume operators had higher rates of arterial thrombosis (0.8% vs 0.4%, p = 0.003), embolism (0.6% vs 0.3%, p<0.001), and major dissection (1.7% vs 0.5%, p<0.001) and lower rates of bleeding events (2.3% vs 3.1%, p = 0.029) compared with lowest volume operators. Across tertiles of operator volume, the rates of in-hospital MALE were comparable among highest and lowest volume operator. Incidence of in-hospital MACE (1.0% vs 1.2%, p = 0.014) and in-hospital mortality (0.7 vs 0.9, p=0.017) were lowest among highest-volume operators as compared with lowest-volume operators (**Table 2**). In the adjusted analysis, undergoing PVI with highest volume operators (vs. lowest volume operators) was associated with lower odds of in-hospital MALE (highest vs lowest-volume: OR 0.72, 95% CI 0.54 – 0.94, p = 0.017) (**Figure 3**). Additionally, intermediate operator volume tertile was also associated with lower risk of MALE as compared to lowest-volume tertile (OR 0.68, 95% CI 0.51 – 0.91, p= 0.008). On further stratification based on PAD categories, this association remained consistent without any statistically significant heterogeneity (p-interaction >0.20). Moreover, no significant association was observed when stratified based on procedural status (elective vs. non-elective). Additional analysis revealed no significant relationship between tertiles of operator volume and MACE (highest-vs lowest-volume: OR 0.71, 95% CI 0.48 – 1.04, p = 0.082). This association remained consistent when stratified based on procedural indication or status.

Female sex, comorbidities such as ESRD on dialysis, coronary artery disease, tobacco use, and acute/chronic limb ischemia were all associated with higher risk of MALE (**Table 3**). Additionally, older age, comorbidities such as hypertension, dialysis, severe lung disease, prior MI, prior HF, and acute/critical limb ischemia were all associated with greater risk of MACE (**Table 4**).

## Discussion

From this analysis of over 60,000 PVI procedures nationwide, there was substantial variability in operator and hospital volume for PVI. In the multivariable adjusted models, incidence of in-hospital MALE was lowest among the highest volume operators but was not associated with tertiles of hospital volume. Conversely, there was no significant relationship between both hospital or operator-volume and MACE events after PVI in the adjusted analysis.

The volume-outcome relationship has been investigated across a wide variety of procedural and surgical domains. In cardiovascular operations such as CABG^13, 16^ and valve surgery^12, 17, 18^, higher hospital- and/or surgeon-volume has been shown to be associated with lower mortality. Similar findings have been seen among vascular surgery procedures such as abdominal aortic aneurysm repair, carotid endarterectomy, and arterial lower limb bypass procedures.^19^ This relationship was also described for some analyses of percutaneous coronary intervention (PCI),^20–23^ transcatheter aortic valve repair (TAVR)^24^ and carotid stenting^25^ where an inverse relationship has been seen with volume and outcomes. However, to our knowledge, the present study is one of few investigations of the volume-outcome relationship in PVI.

Our study aimed to address the knowledge gap by conducting a contemporary assessment of the relationship between hospital and operator volume of PVI and in-hospital outcomes. In this study, highest volume operators were associated with a lower incidence of in-hospital MALE when adjusted for clinical comorbidities and patient demographics. However, hospital-volume was not associated with in-hospital MALE or MACE. These findings seem to be concordant with the “practice makes perfect” hypothesis, which reasons that higher volumes per operator lead to improved decision making, decreased post-operative complications, and operator technique, resulting in improved clinical outcomes. In the past several years, volume has been an appealing quality assessment as it is relatively easy to determine and does not require detailed clinical data. Previous studies have reported significantly better outcomes for vascular procedures with high volume providers, both in the elective and emergent setting.^19^ Similarly, based on our findings, it is conceivable that individual operator volumes may be more significant than institutional volume for optimizing PVI procedural outcomes.

On the contrary, higher hospital volumes were not associated in lowering the incidence of MALE and MACE events. There are many possible reasons for this lack of relationship. As compared with TAVR, which is a more novel technology, PVI procedures utilize similar technology as coronary angioplasty and coronary stenting. Importantly, the relationship between PCI volume and clinical outcomes in contemporary cohorts is much more modest than when the procedure initially was developed,^26, 27^ and perhaps the relationship between any percutaneous procedural volume and outcomes is attenuated with greater temporal procedural familiarity. Consequently, there may be greater familiarity with PVI procedures despite low hospital volumes given crossover of skills from coronary interventions. This may be especially important given that >80% of procedures in the present study occurred in a catheterization laboratory as opposed to with interventional radiology and in an operating room. Moreover, it is not uncommon to have multiple physicians from different specialties with endovascular privileges; Conceivably, these differences in cross specialty practices could lead to variation in care quality and outcomes. Lastly, there are many different PVI procedures, including angioplasty, atherectomy, and stent placement across various lower extremity arteries. Given this heterogeneity in procedures, assessment of general PVI volume may not have enough specificity to properly elucidate the relationship between hospital volume and outcomes. Given these findings, it seems plausible that depending primarily on institutional procedural volume may not adequately discern between low-performing or high-performing institutions in PVI within the contemporary landscape.

These findings have important implications. There is significant variability in performance of PVI by different specialties, with interventional cardiology, interventional radiology, and vascular surgery all performing PVI procedures nationwide.^28^ However, training pathways are heterogenous. The recent ACC/AHA/SCAI guidelines recommended 100 peripheral angiograms and 50 peripheral interventions prior to independent practice, as well as other time-based training competencies across all three specialties.^29^ The present findings are concordant with the above recommendations and highlights the significance of individual operator volumes, potentially outweighing institutional volume in the context of these procedures. Our study findings indicate that operator-volume may be a valuable surrogate to assess PVI quality across these centers.

There are notable limitations to this study. There was limited follow-up data available, and it is not known if longer-term out-of-hospital assessment of both MACE and MALE will alter our findings. Moreover, interpreting MALE rates in patients with CLTI may be challenging since a failed limb salvage procedure requiring amputation would also be classified as a MALE event based on the coding system. Furthermore, the absolute differences in event rates across tertiles is small in our study. The data source utilized was the NCDR PVI registry, which consists of a heterogeneous group of centers across the United States. However, there may be differences between operator specialties and types of centers that participate in the registry and those that do not participate which may have led to residual confounding and may limit generalizability. Additionally, the effects of COVID-19 pandemic in 2020 could have influenced some of the study findings, introducing additional confounding factors that may not have been considered. However, registries are a valuable contributor to real-world evidence, and this manuscript adds to the growing literature regarding the role of volume and its association with clinical outcomes.

## Conclusions

In conclusion, in this analysis of over 60,000 PVI procedures from the NCDR PVI registry, there was substantial variability in operator- and hospital-volume of PVI. After multivariable adjustment, highest volume operators were associated with lower incidence of in-hospital MALE. However, there was no observed association between either hospital-volume of PVI and MACE or MALE. Future studies are warranted, including those with long-term outcome assessment, to evaluate operator and hospital volume as a potential surrogate to assess PVI quality and improve outcomes.

## Data Availability

NCDR database access is available only with special permission

## Abbreviations

PVI: Peripheral vascular interventions
PAD: Peripheral artery disease
MALE: Major adverse limb events
MACE: Major adverse cardiovascular events
NCDR: National Cardiovascular Data Registry
MI: myocardial infarction
ESRD: End stage renal disease
CAD: coronary artery disease
PCI: Percutaneous coronary intervention
CABG: Coronary artery bypass grafting

---

**Central Illustration.**
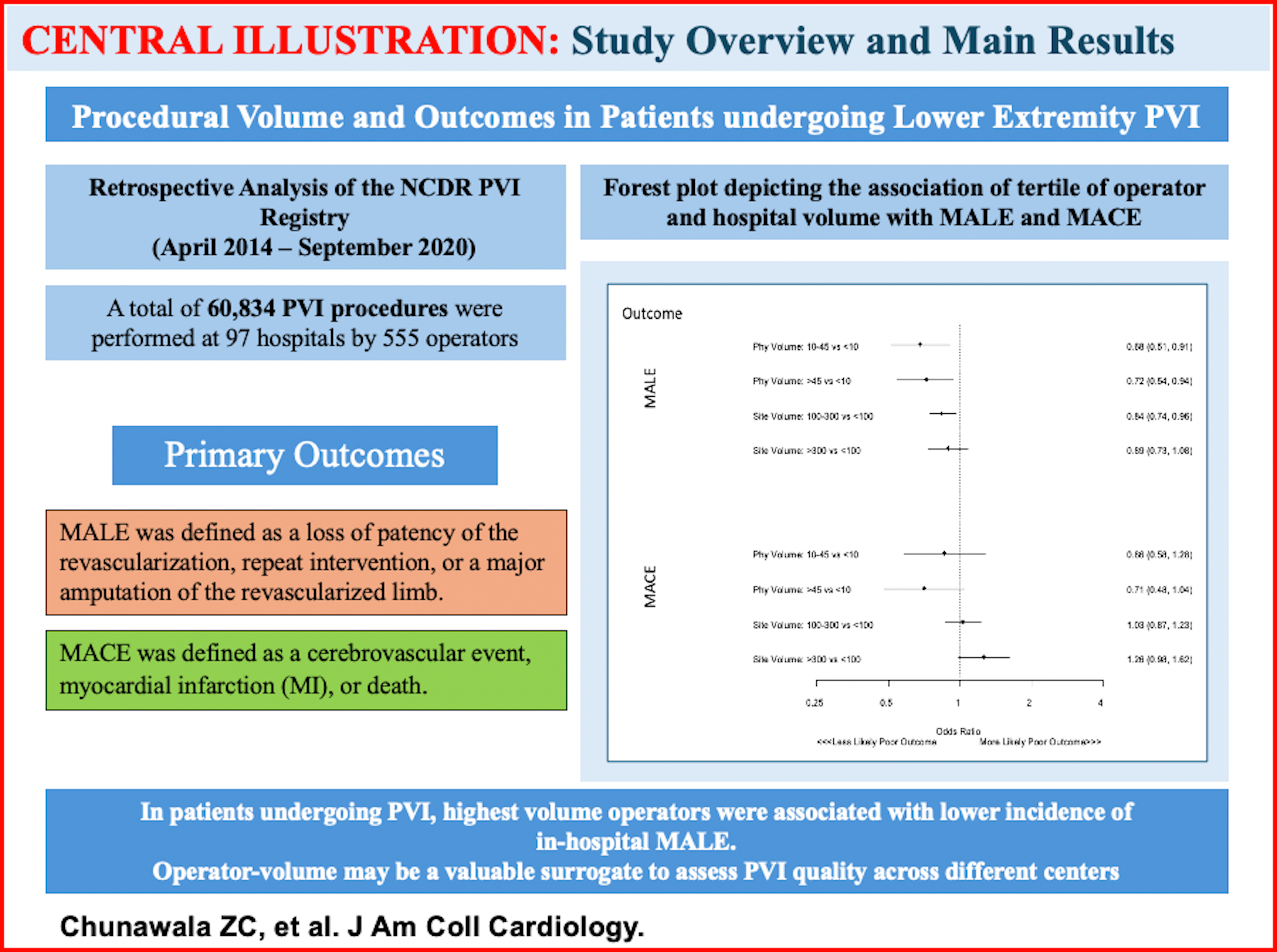
Procedural Volume and Outcomes in patients undergoing lower extremity PVI.

